# Long-term Improvement of Air Quality Associated with Lung Function Benefits in Chinese Young Adults: A Quasi-experiment Cohort Study

**DOI:** 10.1101/2022.01.12.22269121

**Authors:** Shurong Feng, Jiaming Miao, Minghao Wang, Ning Jiang, Siqi Dou, Liu Yang, Yang Ma, Pei Yu, Tingting Ye, Yao Wu, Bo Wen, Peng Lu, Shanshan Li, Yuming Guo

## Abstract

**Background:** Long-term exposure to air pollution is associated with lung function impairment. However, whether long-term improvements in air quality could improve lung function is unclear.

**Methods:** We conducted a prospective quasi-experiment cohort study with 1731 college students in Shandong, China from September 2019 to September 2020, covering COVID-19 lockdown period. Data on PM_2.5_, PM_10_, NO_2_ and SO_2_ concentrations were obtained from China Environmental Monitoring Station. The concentration of O_3_ was obtained from Tracking Air Pollution in China. Lung function indicators included forced vital capacity (FVC), forced expiratory volume in 1 second (FEV1) and forced expiratory flow at 50% of FVC (FEF50%). Linear mixed-effects model was used to examine the associations between the change of air pollutants’ concentrations and the change of lung functions. We also conducted stratified analysis by sex.

**Results:** Compared with 2019, the mean FVC, FEV1 and FEF50% were elevated by 414.4ml, 321.5ml, and 28.4ml respectively in 2020. Every 5μg/m^3^ decrease in annual average PM_2.5_ concentrations was associated with 36.0ml [95% confidence interval (CI):6.0, 66.0ml], 46.1ml (95% CI:16.7, 75.5ml), and 124.2ml/s (95% CI:69.5, 178.9ml/s) increment in the FVC, FEV1, and FEF50%, respectively. Similar associations were found for PM_10_. There was no significant effect difference between male and female.

**Conclusions:** Long-term improvement of air quality can improve lung function among young adults. Stricter policies on improving air quality are needed to protect human health.

**Funding:** Taishan Scholar Program

## Introduction

Air pollution is the fourth leading risk factor for death globally, accounting for 6.67 million deaths in 2019 (Murray et al., 2020). Respiratory system as the direct target organ of air pollution exposure could easily be affected by deleterious air conditions (Schraufnagel et al., 2019). Studies have shown that both short-term and long-term exposures to air pollution could increase the mortality and morbidity of respiratory diseases (Duan, Hao, & Yang, 2020; Liu et al., 2019; Salimi et al., 2018).

Lung function indicators, such as FVC, FEV1 and FEF50% in lung function tests, can be used to reflect the early changes in lung health (Cotes, Chinn, & Miller, 2009; Smith, Emerson, Kurti, Gandhi, & Harms, 2015). Studies have suggested that early adulthood low lung function is associated with the incidence of later respiratory diseases (Agustí, Noell, Brugada, & Faner, 2017; Bui et al., 2018). Current studies suggested the adverse effect of poor air quality on lung function (Panis et al., 2017; Schultz, Litonjua, & Melén, 2017; Zeng et al., 2016), but whether lung function can be improved when exposed to cleaner air is limited. A recent Chinese cohort study has found that for every 10μg/m^3^ reduction in annual average PM_2.5_, the peak expiratory flow (PEF) value could increase by 14.95 L/min for participants at an average age of 60.5 years (Xue et al., 2021). An Switzerland follow-up study of adult with an average age of 41.5 years has found that 10μg/m^3^ reduction in PM_10_ concentrations over the past 11 years could reduce the annual decline rate of FEV1 by 9% (Downs et al., 2007). In California, the Children’s Health Study found that lung-function growth improves with air quality improvements over the study period (Gilliland et al., 2017). These studies are mostly limited to children or old adults, short-term improvement of air quality, or single pollutant exposure design. Individuals with low levels of FEV1 and FVC in young adults period were at increased risk for early pulmonary mortality (Vasquez, Zhou, Hu, Martinez, & Guerra, 2017). However, few studies have investigated the association between long-term improvement of air quality and changes of lung function in young adults.

In 2020, after the COVID-19 outbreak, to prevent the spread of the pandemic diseases, China has implemented large-scale quarantine strategies during January 20 to April 8, 2020 (S. Chen, Yang, Yang, Wang, & Bärnighausen, 2020; Fan, Zhan, Yang, Liu, & Zhan, 2020; Tian et al., 2020). Factories and the public transport network emissions were reduced (Wang et al., 2020). The air pollutants concentrations during the COVID-19 period decreased significantly in comparison with the same time period of the previous year (Nie et al., 2021). This provides a good opportunity to explore the effects of long-term improvement of air quality on lung function.

Therefore, we extracted data from the Chinese Undergraduates Cohort to evaluate the association between air quality improvement and changes of lung function among young adults. For better assessing the differences of the effects between sex, stratified analysis was also conducted.

## Materials and Methods

### Study design and study population

Participants were selected from the Chinese Undergraduate Cohort (CUC). The CUC used random cluster sampling method to choose study subjects. The CUC was established to explore the impact of lifetime environmental (such as air pollution, ambient temperature, surrounding greenness) exposure and behavior on health outcomes (physical and mental health). In 2019, we recruited from the CUC 2,192 participants from Binzhou Medical University in Yantai, Shandong Province. They were all graduated high school in Shandong Province. Before the lung function test, participants completed the survey including general questions on demographics, lifestyle and disease history, etc. Finally, in total 1,731 participants with complete results of spirometry and questionnaire information in both 2019 and 2020 were included in the analyses (Fig.1). Long-term exposure was estimated as the annual average value of the pollutant concentration one year before the individual’s lung function test. We matched the exposure concentration based on the participants’ time at school and home. In 2020, due to the impact of COVID-19, participants stay at home from January 10, 2020 to August 22, 2020.

**Fig. 1.**
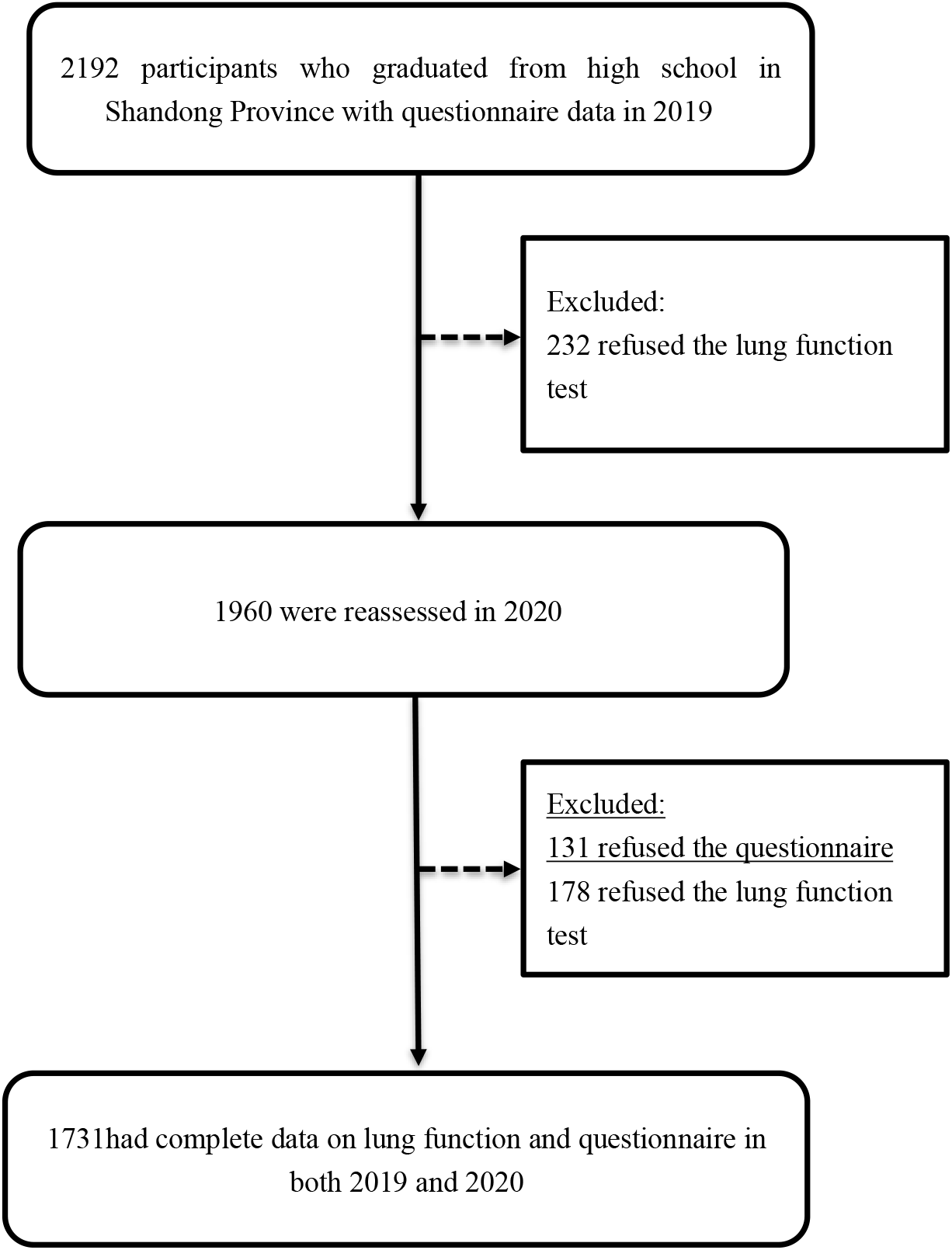
Flow chart of cohort study participants.

### Ethical approval

The study was approved by the ethics committee of Binzhou Medical College, and the students’ informed consent was obtained before participating.

### Lung function test

Participants in this study did two-round lung function tests. The first lung function test was performed between September 1 and September 29, 2019. The follow-up test was conducted from September 5 to November 15, 2020. The two-year spirometry were assessed indoor by Gest HI - 101 spirometer (Chest, Tokyo, Japan). Lung function tests were performed according to the European Respiratory Society specifications (Brusasco et al., 2005). A complete lung function test contains three indicators: slow vital capacity (SVC), FVC, and maximum voluntary ventilation (MVV). Specific quality controls were performed as follows: (1) all tests were done by well-trained professionals; (2) spirometers were calibrated before daily lung function test, and immediate calibration check after replacing; (3) no blocked mouthpiece, air leakage, early termination or cut-off of expiration any detector; (4) extrapolation volume less than 150 mL or 5% of the FVC, and a forced expiratory time exceeding 6 s; (5) expiratory platform in volume-time curve and reproducible test with three acceptable flow-volume curves. We selected the best measurement results of the participants as their lung function results (Sylvester et al., 2020). Finally three pulmonary function indices: FVC, FEV1 and FEF50% were included in present study.

### Exposure

We obtained the daily average concentrations of PM_2.5_, PM_10_, NO_2_ and SO_2_ for 16 cities in Shandong Province from China National Environmental Monitoring Center (http://www.cnemc.cn/). The daily average concentrations data at the city-level was calculated based on the average of the data from nation-level monitoring stations of the 16 cities. The number of nation-level monitoring stations in each city was shown in Appendix 1—Table 1. All monitoring stations are not located close to traffic, industrial pollution or other local pollution sources, nor can be affected by emissions from buildings or large houses. We used Tracking Air Pollution in China (http://tapdata.org.cn) shared daily average 10km×10km maximum 8-hour O_3_ grid data for 16 cities from 2018 to 2020. This O_3_ estimation dataset was generated by a data-fusion algorithm that combines in situ observations, satellite remote sensing measurements, and model results from the community multiscale air quality model (Xue et al., 2020). Participants’ living addresses and school addresses were obtained from the questionnaire. The home and school addresses of each participant were coded into city code to match with the exposure concentrations in each city. We matched the daily PM_2.5_, PM_10_, SO_2_ and NO_2_ obtained from the monitoring station according to the participants’ time at school and home. And matched the nearest grid data of O_3_ according to the latitude and longitude of the participant’s home or school. Estimated the annual average exposure concentration based on the daily exposure concentration.

### Statistical methods

We used linear mixed-effects models for longitudinal data analysis to examine associations between annual average air pollution exposure and lung function (Brown, 2021). The mixed model included city and subject as random effect parameters and adjusted potential confounders based on associations with air pollution exposure and lung function. The mixed model adjusted for sex, age, body mass index (BMI), lifestyle factors (smoking status, alcohol drinking, physical activity), respiratory disease history, home location (rural or urban areas), socioeconomic status, temperature, and relative humidity in the corresponding period. The respiratory diseases history included the diagnosis of the following diseases: allergic rhinitis, asthma, chronic bronchitis, pneumonia, emphysema, tuberculosis and other lung diseases. The socioeconomic status was categorized into advantage and disadvantage groups according to the annual family income cutoff of 50,000RMB (≈US$7730.61) (Martinson, Chang, Han, & Wen, 2018). To control for the non-linear effects of temperature and relative humidity, we added a natural cubic spline term for them. To facilitate the comparison of estimates between pollutants, effect estimates of the milliliters change in lung function were presented as per 5µg/m^3^ increment in air pollutant concentrations. We repeated analyses using data stratified by sex (male and female) to explore potential effect modifications by them. To adjust the potential confounding effects of co-pollutants, we ran two pollutant models. Since PM_2.5_ and PM_10_ are highly correlated, these two are not included in the same model.

We conducted the following sensitivity analyses: the air pollution exposure duration of 10 and 11 months before lung function measurement were used to assess the robustness of our results. All analyses were conducted in R software, with the “nlme” package (version 4.0.3). The threshold of statistical significance was p value of < 0.05 for 2-sided test.

## Results

### Subject’s characteristics

Table 1 shows the demographic characteristics and lung function test results. There were no significant changes in demographic characteristics in 2019 and 2020. This was a young adult cohort with a small proportion of smokers and a slight majority of females. There were 131 (7.5%) subjects with self-reported respiratory disease history. The mean FVC was 2733.9 ml, FEV1 was 2668.3 ml and FEF50% was 4601.1 ml/s in 2019. In 2020, the mean FVC, FEV1 and FEF50% was 3148.0ml, 2989.8ml and 4629.5ml/s, respectively. Overall, the mean value of FVC, FEV1 and FEF50% in 2020 was higher than that in 2019. Appendix 1—Table 2 shows the demographic baseline characteristic differences between participants and those who did not participate in the study. Compared with the nonparticipants, there were more women among the participants (by 8.2%, Appendix 1—Table 2). The geographical distribution of the participants’ home addresses is shown in 10. Appendix 1—figure 1.

**Table 1.** Demographic characteristics and lung function outcomes among participants (N = 1731).

| Participant characteristics | Year |  |
| --- | --- | --- |
|  | 2019 | 2020 |
| Demographic characteristic |  |  |
| Age (years) | 18.2±0.6 | 19.0±0.6 |
| Female (%) | 1030 (59.5) | 1030 (59.5) |
| Urban (%) | 886 (51.1) | 886 (51.1) |
| BMI | 22.2±6.1 | 21.8±5.7 |
| Cigarette smoke exposure (%) |  |  |
| Smoking status | 30 (1.7) | 28 (1.6) |
| Passive smoking | 230 (13.2) | 222 (12.8) |
| Alcohol consumption | 85 (4.9) | 87 (5.0) |
| Physical activity at least once a week (%) |  |  |
| Yes | 1325 (76.5) | 1349 (77.9) |
| No | 406 (23.5) | 382 (22.1) |
| Lung disease history (%) |  |  |
| Yes | 318 (18.4) | 159 (9.2) |
| No | 1413 (81.6) | 1572 (90.8) |
| Socioeconomic status (%) |  |  |
| Socioeconomic-advantage | 680 (39.2) | 680 (38.0) |
| Socioeconomic-disadvantage | 1051 (60.7) | 1051 (61.9) |
| Lung function index (ml) |  |  |
| FVC | 2733.9±827.6 | 3148.0±775.9 |
| FEV1.0 | 2668.3±780.7 | 2989.8±695.2 |
| FEF50% | 4601.1±1371.4 | 4629.5±1330.1 |

### Air pollution exposure

Fig. 2 shows the distribution of pollutant concentrations in 2019 and 2020. The average pollutant concentrations in 2020 were lower than in 2019. The annual pollutant concentrations of each city are shown in Figure 2–Source Data 1. The differences in the annual average pollutant concentrations between 2019 and 2020 are shown in Figure 2–Source Data 2. We found that the annual average concentrations of PM_2.5_ decreased from 50.38μg/m^3^ in 2019 to 46.86μg/m^3^ in 2020, PM_10_ from 96.67μg/m^3^ to 85.25μg/m^3^, SO_2_ from 14.72μg/m^3^ to 11.68μg/m^3^, NO_2_ from 36.00μg/m^3^ to 31.10μg/m^3^, and O_3_ from 109.99μg/m^3^ to 107.97μg/m^3^.

**Fig. 2.**
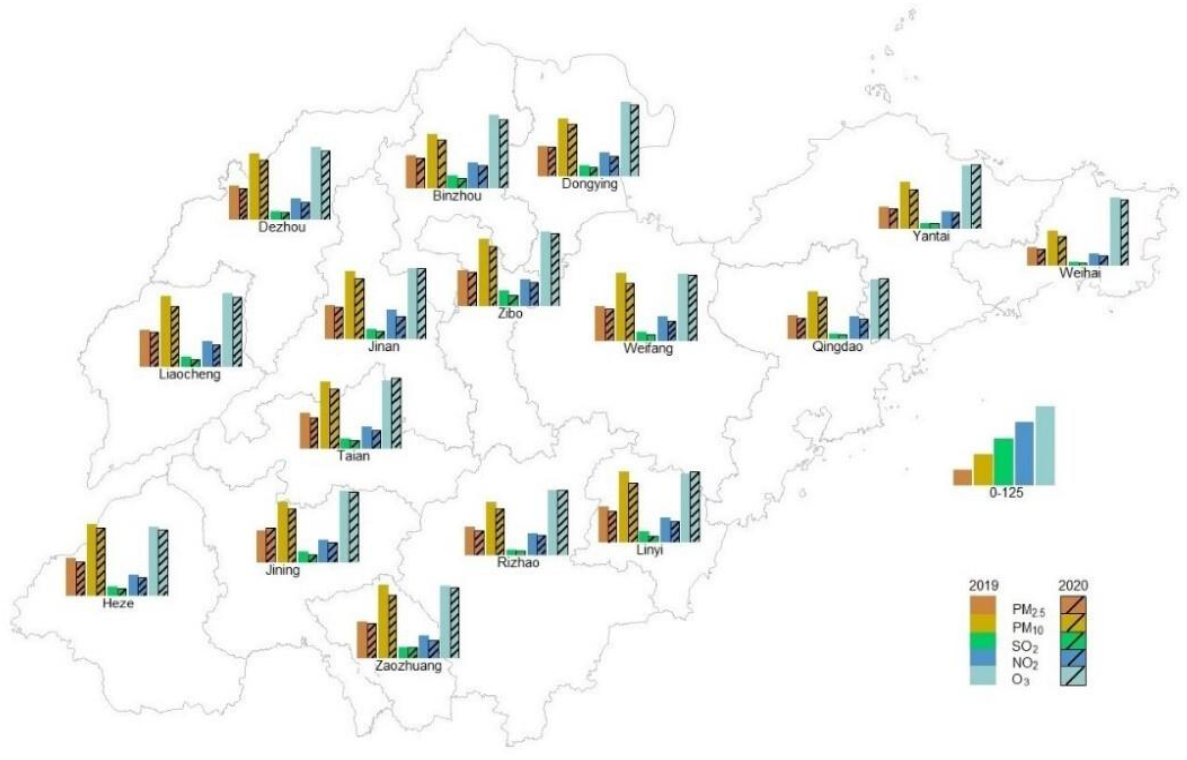
The annual average pollutant concentrations in 16 cities in 2019 and in 2020 (μg/m^3^). The height represents the value of air pollutant concentration. (Figure 2–Source Data 1, Figure 2–Source Data 2)

### Association between lung function and air pollutants

Fig. 3 shows the association between long-term exposure to pollution and lung function. Long-term air pollutants improvement were significantly associated with the increment of FVC, FEV1, and FEF50% levels. Each 5μg/m^3^ reduction of PM_2.5_ was associated with 36.0ml [95% confidence interval (CI):6.0, 66.0ml], 46.1ml (95% CI:16.7, 75.5ml), and 124.2ml/s (95% CI:69.5, 178.9ml/s) increment in the FVC, FEV1, and FEF50%, respectively. The same trend was found in PM_10_, each 5μg/m^3^ reduction in PM_10_ was associated with 17.9ml (95% CI: 1.4,34.4ml), 17.9ml (95% CI: 1.3,34.5ml), and 56.3ml/s (95% CI: 23.4,89.1ml/s) increment in FVC, FEV1, and FEF50%, respectively. We also found the increment of 193.4ml/s (95% CI: 46.3,340.5ml/s) for FEF50% with each 5μg/m^3^ reduction in NO_2_.

**Fig. 3.**
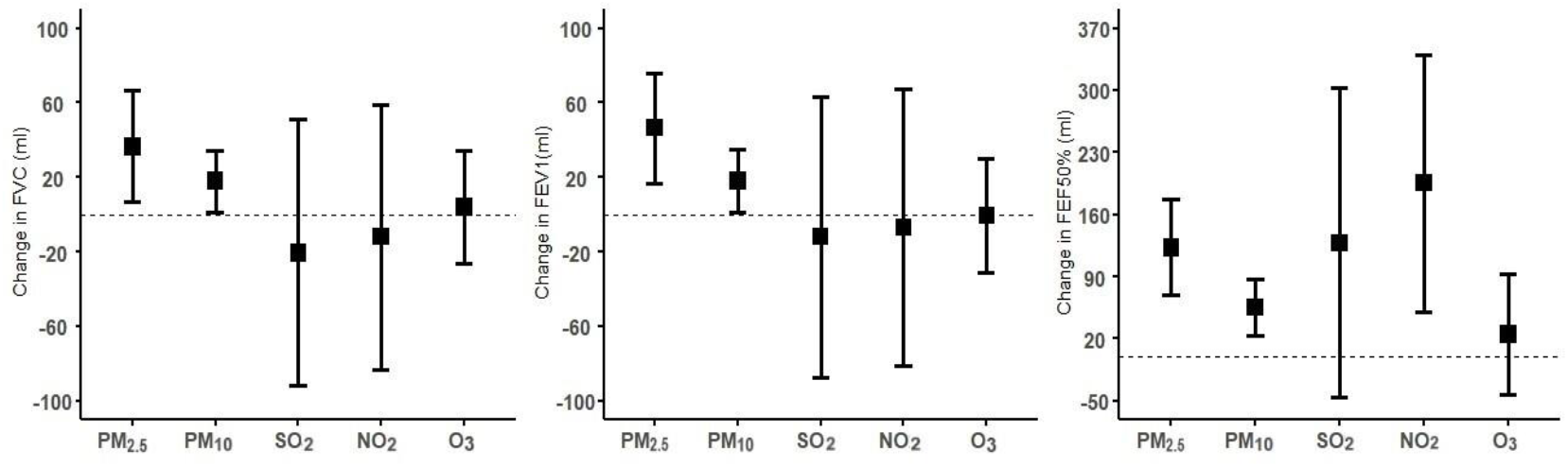
The association between 5μg/m^3^ decrease in the average annual pollutant concentrations and changes in lung function parameters. The linear mixed models included city and subject as random effect and adjusted for ambient air pollutants, sex, age, BMI, lifestyle factors (smoking status, alcohol drinking, physical activity), respiratory disease history, home location (rural or urban areas), socioeconomic status, temperature, relative humidity, wind speed. (Figure 3–Source Data 1)

Fig. 4 shows the results of the two-pollutant model. The two models showed that the inclusion of NO_2_, SO_2_ and O_3_ in the particulate matter model slightly improved effect estimates of PM_2.5_ and PM_10_ on FVC and FEV1, especially after adjusting for NO_2_. Including particulate matter in the NO_2_, SO_2_ and O_3_ models did not change the coefficient estimates of lung function.

**Fig. 4.**
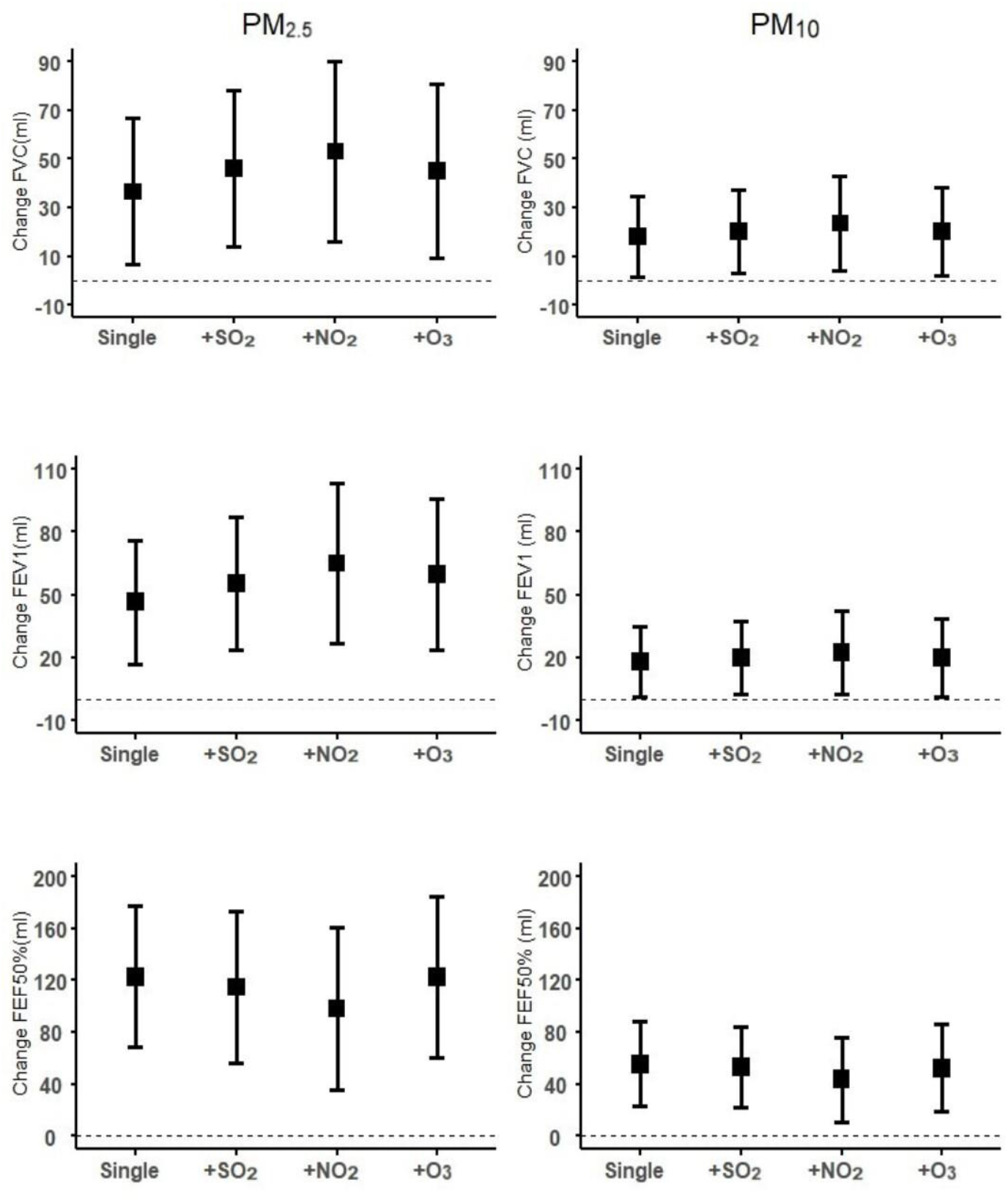
The association between 5μg/m^3^ decrease in the average annual pollutant concentrations and changes in lung function parameters in single and two-pollutant models. (Figure 4–Source Data 1)

### Stratified and sensitivity analysis

Appendix 1—Table 3 shows the stratified analysis results. We did not observe significant effect modification by sex on associations with FVC, FEV1, or FEF50%.

The results of sensitivity analyses were presented in Appendix 1—Table 4-5. The estimation was robust when adjusting the pollutant exposure time to 10 and 11 months before the lung function test.

## Discussion

Our quasi-experiment cohort study found that reductions in PM_2.5_ and PM_10_ concentrations were significantly associated with the increment in FVC, FEV1 and FEF50% among young adults, after controlling for the physiological increase in lung function caused by age in the model. Among all the outcome variables, the small airway index FEF50% was more sensitive to a variety of pollutants, and the estimated change was the largest. After adjusting for gaseous pollutants, the PM impact was estimated to be stronger. We did not observe significant effect modification by sex on associations between air pollution and lung function.

Air pollution has always been a global concern. In high-income countries, studies have explored the benefits of air pollution improvement on lung function. In California, researchers used longitudinal data on 2,120 participants from three cohorts (1993-2001; 1997-2004; 2007-2011) aged 11 to 15 to study the effects of policy-driven air pollution interventions on children’s lung development, the study showed that if the PM_2.5_ concentration had been reduced by 30% from the levels observed in the 1993 cohort, the growth of FVC and FEV1 would be increase by 4.5% and 2.5% compared with observed growth, respectively. Similar results were also observed for NO_2_ (Urman et al., 2020). Our results also indicated that the decline in PM_2.5_ exposure was associated with the increment in FVC, FEV1 and FEF50%. A panel study of the long-term effects of personal PM_2.5_ exposure on lung function conducted in Wuhan and Zhuhai, China showed that subjects with persistently high levels of personal PM_2.5_ exposure decreased their FEV1/FVC by an additional 3.63% within three years, and a decrease of 7.15% in six years compared with subjects with persistently low exposure levels (Zhou et al., 2020). All studies have shown that reducing exposure to air pollution was essential for improving lung function.

Our research shows that when air quality improved, the effects of PM_2.5_ and PM_10_ levels reduction on lung function increment were slightly stronger at FEV1 than FVC. FEV1 is a common indicator for obstructive ventilation disorders (Barreiro & Perillo, 2004). Our results supported that obstructive disease benefits more than restrictive diseases. The findings on the associations between long-term air pollution exposure and lung function indicators were limited and inconsistent. A longitudinal study of adults between 18 and 60 years old in Switzerland found that the reduction of PM_10_ in 11 years was associated with a slower decline in FEV1 but not FVC (Downs et al., 2007), indicating an obstructive disease beneficial effect. A Normative Aging Study cohort in the Boston found that the cross-sectional effects of black carbon (a traffic-related constituent of particulate matter weighted toward diesel) were a bit stronger on FEV1 compared with FVC, suggesting an obstructive effect. However, the longitudinal effects were slightly stronger on FVC compared with FEV1 (Lepeule et al., 2014). A cohort study of 11 to 15 years children in California found that if PM_2.5_ and NO_2_ concentrations were lower than actually observed by 10%, 20% and 30%, respectively, children’s lung function would increase and FVC would increase more than FEV1 (Urman et al., 2020). The discrepancy in these studies may be attributable to different study subjects and exposure patterns. Additional research is needed to examine whether long-term air quality improvements are more beneficial to obstructive diseases or restrictive diseases or whether air quality improvements have different age effects.

Few studies have examined the effects of long-term air pollution exposure on FEF50%. FEF50% is the instantaneous flow rate when 50% of the force is exhaled. Studies have shown that FEF50% is a feasible parameter for identifying small airway dysfunction early (Yuan et al., 2019), FEF50% predicted as the best parameter to predict absence of airway hyperresponsiveness (Peled et al., 2021). Our study found that FEF50% was extremely sensitive to air pollutants. The decrease of PM_2.5_, PM_10_ and NO_2_ by 5μg/m^3^ were associated with an increment of 124.2ml/s, 56.3ml/s and 193.4ml/s respectively. The impact of various pollutants on FEF50% was slightly greater than the impact on FEV1 and FVC. This finding suggests that air quality control may have more benefits on small airway functions than on large airway functions.

We conducted a stratification analysis by sex. In our study, we did not observe significant effect modification by sex on associations between air pollution and lung function. Previous literature showed that men were more sensitive to air pollution (Schultz et al., 2012), while some studies reported that women were more sensitive to air pollution (C. Chen et al., 2018). One possible explanation for the sex difference is that men are relatively active and spend more time outdoors than women (He et al., 2010). Men tend to have more extensive exposures. In our study, outdoor activities had been constrained due to the impact of COVID-19, so the difference in outdoor exposure between men and women decreased. This could partly explain our insignificant sex modification effects.

People were rarely exposed to a single air pollutant. Recent studies have proposed that air quality management must evolve from the perspective of a single pollutant to a multi-pollutant approach for emission monitoring (Dominici, Peng, Barr, & Bell, 2010; Huang, Rappold, Graff, Ghio, & Devlin, 2012). In two pollutant models, we found that inclusion of NO_2_, SO_2_ and O3 in the particulate matter model slightly improved effect estimates of PM_2.5_ and PM_10_ on FVC and FEV1, especially after adjusting for NO_2_. In consistent with our results, a longitudinal study of primary school children in China (C. Chen et al., 2018), found that adjusting for NO_2_ or SO_2_ can increase the association between particulates and lung function. Animal experiments that co-exposure to air pollution increased the expression of interleukin-6 (IL-6) and tumor necrosis factor-α (TNF-a) were demonstrated (Zhang, Ji, Ku, & Sang, 2016), and further promotes the inflammatory response of the trachea. In addition, as a carrier of irritant gases, PM has the ability to absorb and transport NO_2_ into the trachea, consequently resulting in greater harm to the trachea (Boren, 1964). Therefore, air pollution control changed from a single pollutant to multiple pollutants can benefit more for the public.

The study had several strengths. First, prospective cohort study design allowed us to explore the causal relationship between long-term air pollution exposure and lung function. Secondly, the natural decline of the air pollutants concentrations offered us a good opportunity to conduct a quasi-experiment cohort to clarify the health benefits of air quality improvements.

Our study had some limitations. First, due to the availability of data, except O_3_, we used data from city-level monitoring stations as personal exposure concentrations. Second, although we adjusted the impact of potential confounding factors, there are still some unmeasured confounding factors that we cannot rule out. Third, a potential source of bias in all cohort studies is the loss to follow-up. In our study, there is a certain difference in sex distribution between non-participants and participants, but the stratified analysis of sex found that there was no significant effect modification by sex on associations between air pollution and lung function.

## Conclusions

Long-term improvements in ambient air quality, such as PM_2.5_ and PM_10_, can increase lung function, especially FEF50% among young adults. Given the number of health outcomes associated with impaired lung function, such as COPD and cardiovascular disease, it is vital to improve air quality.

## Data Availability

Investigators and potential collaborators interested in the datasets will be asked to submit a brief concept note and analysis plan. Requests will be vetted by PHD. Yuming Guo and Peng Lu and appropriate datasets will be provided through a password protected secure FTPS link. Recipients of study data will be asked to sign a data sharing agreement that specifies what the data may be used for (specific analyses), criteria for acknowledging the source of the data, and the conditions for publication.

## Acknowledgements

The authors are grateful to all students of Binzhou Medical College who participated in this study.

## Competing Interest Statement

The authors have declared no competing interest.

## Data availability

Data will be shared under the auspices of the Principal Investigators. Investigators and potential collaborators interested in the datasets will be asked to submit a brief concept note and analysis plan. Requests will be vetted by PHD. Yuming Guo and Peng lu and appropriate datasets will be provided through a password protected secure FTPS link. No personal identifying information will be made available to any investigator. General de-identified datasets will be prepared that can accommodate the majority of requests. These will be prepared, with documentation, as the data is cleaned for analysis in order to reduce time and resources required to respond to individual requests. Recipients of study data will be asked to sign a data sharing agreement that specifies what the data may be used for (specific analyses), criteria for acknowledging the source of the data, and the conditions for publication. It will also stipulate that the recipient may not share the data with other investigators. Requests for data use must be made directly to the PI and not through third parties.

---

**Figure 2–Source Data 1.**
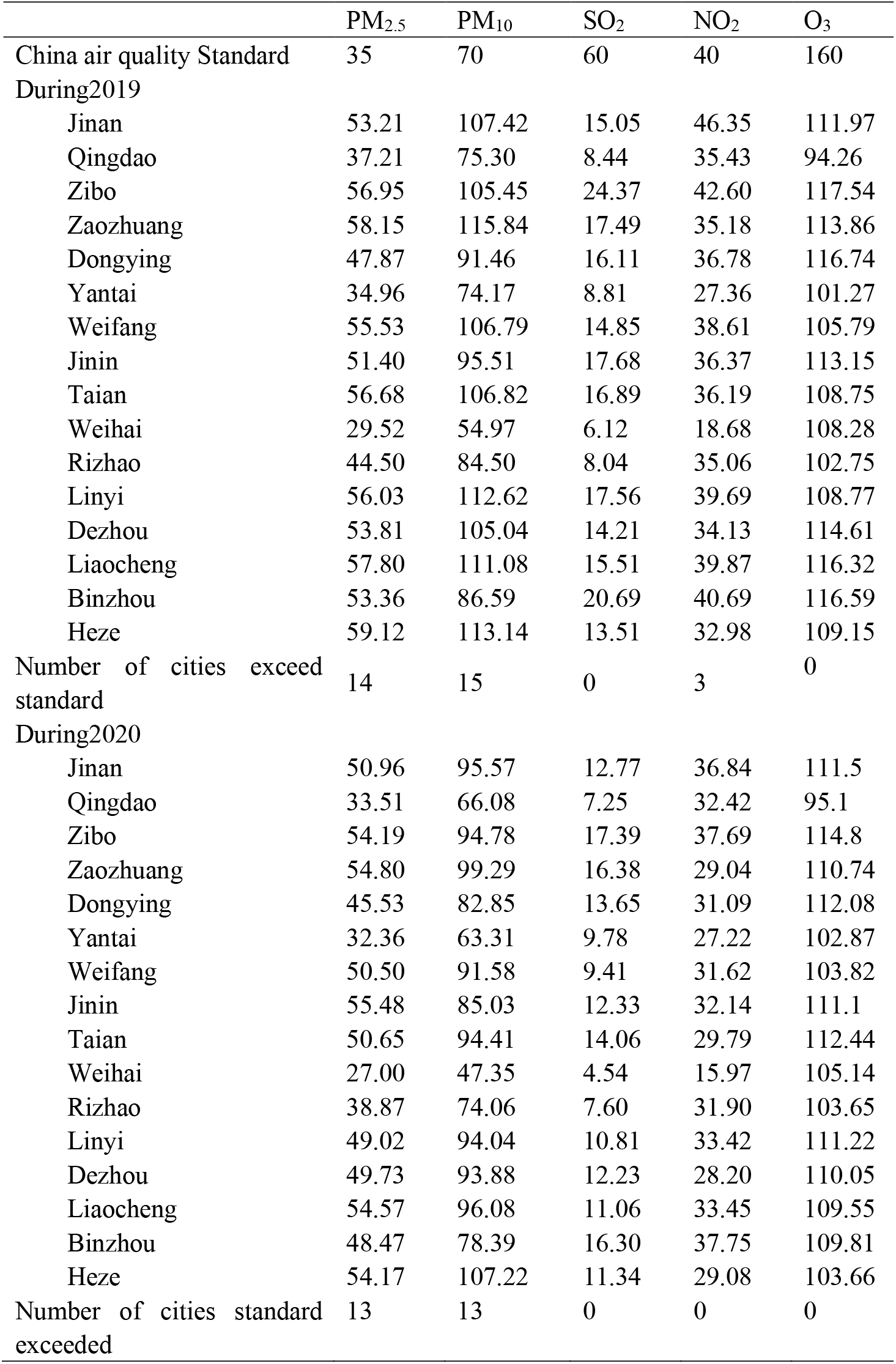
Average annual pollutant concentration in 16 cities (μg/m^3^)

**Figure 2–Source Data 2.**
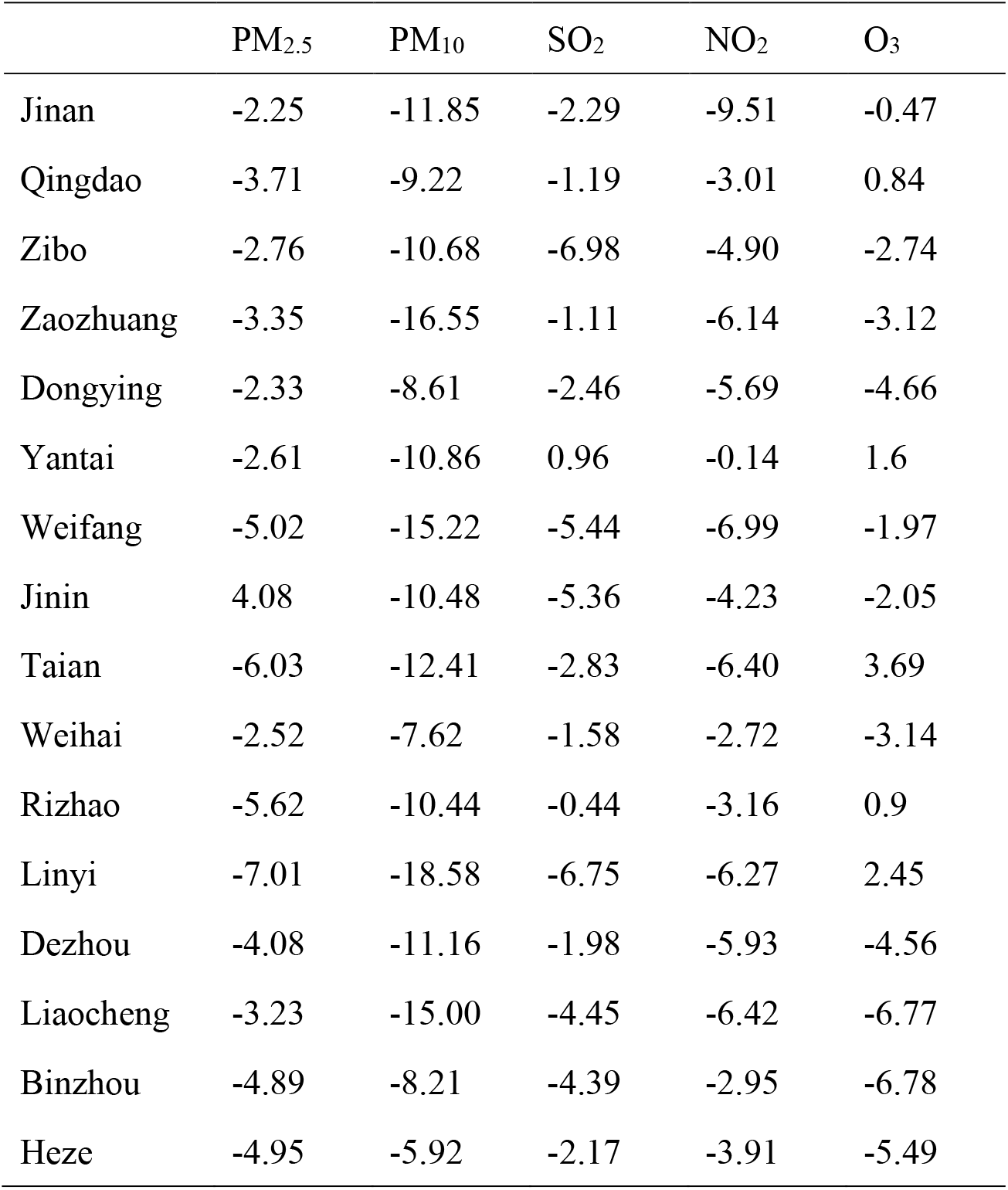
Differences in annual average pollutant concentrations in 16 cities in 2 years(μg/m^3^)

**Figure 3–Source Data 1.**
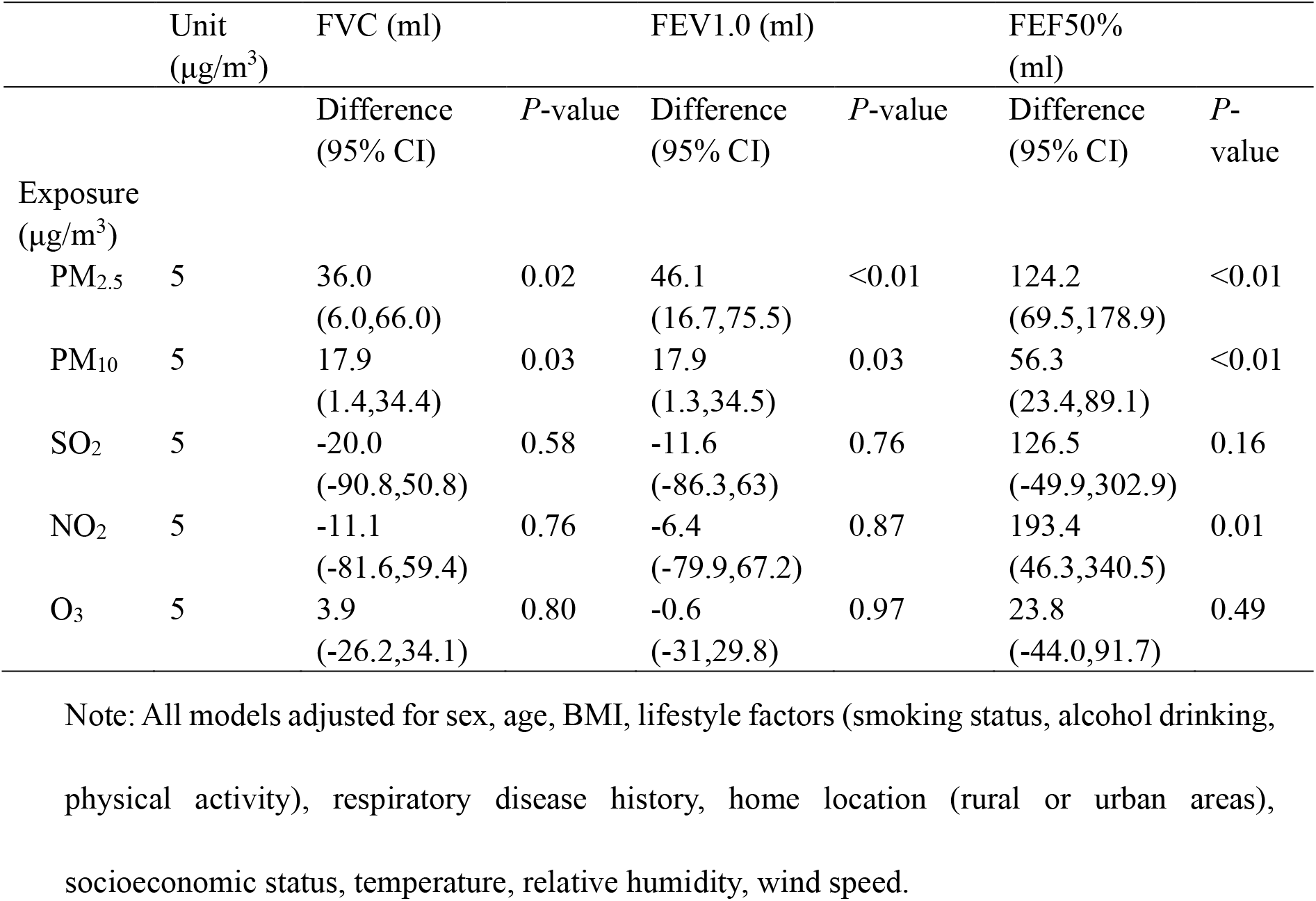
Association between reduction in concentrations of air pollutants and the growth of lung function in young adults

**Figure 4–Source Data 1.**
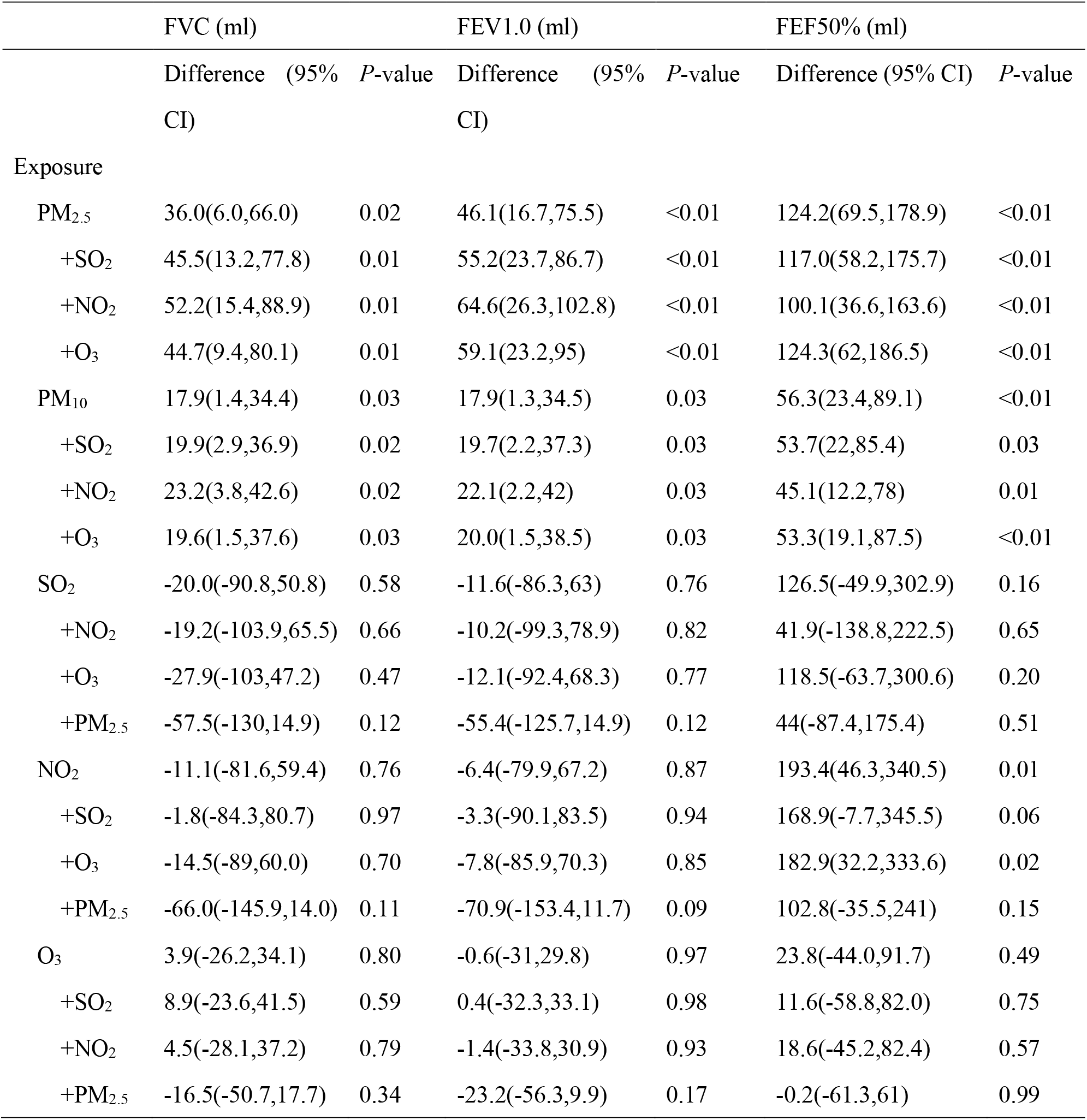
The association between 5μg/m^3^ decrease in the average annual pollutant concentrations and lung function parameters of the two-pollutant model

## Appendix 1

**Appendix 1—figure 1.**
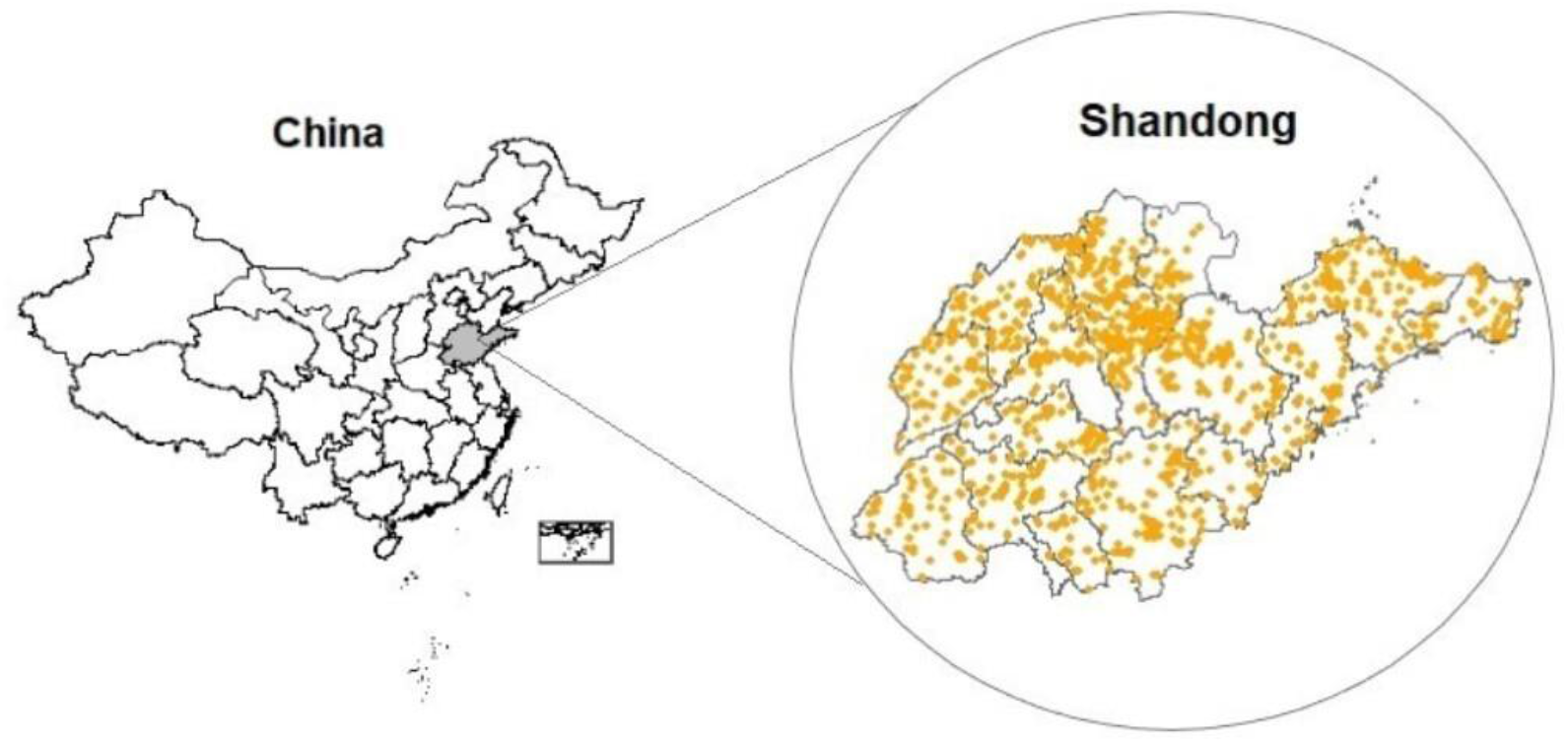
Geographical distribution of the study population Note: The gray area on the left represents Shandong Province; The yellow dots on the right represent participants’ home addresses.

**Appendix 1—Table 1.** Number of control-level environmental monitoring stations in 16 cities in Shandong Province during the study period

| City | Number of stations | City | Number of stations |
| --- | --- | --- | --- |
| Jinan | 8 | Taian | 3 |
| Qingdao | 9 | Weihai | 3 |
| Zibo | 6 | Rizhao | 3 |
| Zaozhuang | 5 | Linyi | 4 |
| Dongying | 4 | Dezhou | 3 |
| Yantai | 6 | Liaocheng | 3 |
| Weifang | 5 | Binzhou | 3 |
| Jinin | 3 | Heze | 3 |

**Appendix 1—Table 2.** Baseline characteristics of the present study participants and participants who did not participate in this study

|  | Participants | Nonparticipants | *P*-value |
| --- | --- | --- | --- |
|  | (N = 1731) | (N = 461) |  |
| Age (years) | 18.2±0.6 | 18.1±1.4 | 0.52 |
| Female (%) | 1030 (59.5) | 235 (51.0) | <0.01 |
| Urban (%) | 886 (51.1) | 238 (51.6) | 0.87 |
| BMI | 22.2±6.1 | 22.6±5.2 | 0.90 |
| Cigarette smoke exposure (%) |  |  |  |
| Smoking status | 30 (1.7) | 6 (1.3) | 0.52 |
| Passive smoking | 230 (13.2) | 68 (14.8) | 0.42 |
| Alcohol consumption | 85 (4.9) | 33 (7.2) | 0.06 |
| Physical activity at least once a week (%) |  |  |  |
| Yes | 1325 (76.5) | 362 (78.5) |  |
| No | 406 (23.5) | 99 (21.5) | 0.37 |
| Lung disease history (%) |  |  |  |
| Have | 318 (18.4) | 83 (18.0) |  |
| No have | 1413 (81.6) | 378 (82.0) | 0.86 |
| Socioeconomic status (%) |  |  |  |
| Socioeconomic-advantage | 680 (39.2) | 196 (42.5) |  |
| Socioeconomic-disadvantage | 1051 (60.7) | 265 (57.5) | 0.23 |

**Appendix 1—Table 3.** The association between 5μg/m^3^ decrease in the average annual pollutant concentrations and changes in lung function parameters stratified by sex

| Lung<br>function<br>(ml) | Pollutants | Males |  | Females |  |
| --- | --- | --- | --- | --- | --- |
|  |  | Difference (95%<br>CI) | *P*-<br>value | Difference (95%<br>CI) | *P*-<br>value |
| FVC | PM<sub>2.5</sub> | 41.8(-15.3,99) | 0.15 | 23.3(-10.0,56.7) | 0.17 |
|  | PM<sub>10</sub> | 15.0(-15.6,45.7) | 0.34 | 12.0(-6.5,30.4) | 0.21 |
|  | SO<sub>2</sub> | -51.0(-99.3, -2.7) | 0.04 | -53.5(-129.5,22.5) | 0.17 |
|  | NO<sub>2</sub> | -66.9(-200.5,66.8) | 0.33 | 18.0(-55.3,91.3) | 0.63 |
|  | O<sub>3</sub> | -14.6(-71.1,41.9) | 0.61 | 14.1(-18.9,47.1) | 0.40 |
| FEV1 | PM<sub>2.5</sub> | 57.0(-2.0,116.0) | 0.06 | 31.6(0.6,62.6) | 0.05 |
|  | PM<sub>10</sub> | 10.8(-21.2,42.9) | 0.51 | 13(-4.2,30.2) | 0.14 |
|  | SO<sub>2</sub> | -45.9(-96.6,4.9) | 0.08 | 55.6(-15,126.3) | 0.12 |
|  | NO<sub>2</sub> | -23.1(-77.8,31.5) | 0.41 | 23.3(-44.8,91.4) | 0.50 |
|  | O<sub>3</sub> | -21.7(-77.9,34.4) | 0.45 | 16.4(-14.2,47.1) | 0.29 |
| FEF50% | PM<sub>2.5</sub> | 135.8(29.9,241.8) | 0.01 | 99.7(38.1,161.3) | <0.01 |
|  | PM<sub>10</sub> | 63.3(1.6,125.1) | 0.04 | 44.4(10.3,78.5) | 0.01 |
|  | SO<sub>2</sub> | 123.2(-152,398.5) | 0.38 | 134.3(-24.2,292.8) | 0.10 |
|  | NO<sub>2</sub> | 261.6(11.9,511.3) | 0.04 | 161.5(26.6,296.4) | 0.02 |
|  | O<sub>3</sub> | 43.0(-71,156.9) | 0.46 | 49.4(-12.6,111.4) | 0.12 |

**Appendix 1—Table 4.** The association between 5μg/m^3^ decrease in the average annual pollutant concentrations in 11 months before the lung function measurement and lung function parameters

|  | FVC (ml) |  |  | FEV1.0 (ml) |  |  | FEF50% (ml) |  |  |
| --- | --- | --- | --- | --- | --- | --- | --- | --- | --- |
|  | Difference | (95% | *P*-value | Difference | (95% | *P*-value | Difference | (95% | *P*-value |
|  | CI) |  |  | CI) |  |  | CI) |  |  |
| Exposure |  |  |  |  |  |  |  |  |  |
| PM<sub>2.5</sub> | 30.0(3.1,56.9) |  | 0.03 | 39.0(12.2,65.8) |  | <0.01 | 102.5(54.1,150.9) |  | <0.01 |
| PM<sub>10</sub> | 17.5(1.9,33.1) |  | 0.03 | 17.5(1.5,33.5) |  | 0.03 | 34.0(1.1,66.9) |  | 0.04 |
| SO<sub>2</sub> | -16.0(-88.1,56.1) |  | 0.67 | -3.0(-80.0,0.74) |  | 0.94 | 148.0(-23.2,319.2) |  | 0.09 |
| NO<sub>2</sub> | -4.5(-70.4,61.4) |  | 0.90 | -4.5(-74.0,65.0) |  | 0.90 | 123.5(-16.6,263.6) |  | 0.08 |
| O<sub>3</sub> | 2.0(-27.6,31.6) |  | 0.88 | -1.0(-31.0,29.0) |  | 0.96 | 37.5(-26.9,101.9) |  | 0.25 |

**Appendix 1—Table 5.** The association between 5μg/m^3^ decrease in the average annual pollutant concentrations in 10 months before the lung function measurement and changes in lung function parameters

|  | FVC (ml) |  |  | FEV1.0 (ml) |  |  | FEF50% (ml) |  |  |
| --- | --- | --- | --- | --- | --- | --- | --- | --- | --- |
|  | Difference | (95% | *P*-value | Difference | (95% | *P*-value | Difference | (95% | *P*-value |
|  | CI) |  |  | CI) |  |  | CI) |  |  |
| Exposure |  |  |  |  |  |  |  |  |  |
| PM<sub>2.5</sub> | 40.0(13.7,66.3) |  | <0.01 | 47.0(21.8,72.2) |  | <0.01 | 101.0(53.8,148.2) |  | <0.01 |
| PM<sub>10</sub> | 16.5(1.3,31.7) |  | 0.03 | 17.0(1.4,32.6) |  | 0.03 | 33.5(1.7,65.3) |  | 0.04 |
| SO<sub>2</sub> | 15.5(-61.0,92.0) |  | 0.69 | 30.0(-50.2,110.2) |  | 0.46 | 169.0(2.4,335.6) |  | 0.05 |
| NO<sub>2</sub> | 9.5(-59.4,78.4) |  | 0.79 | 10.5(-61.3,82.3) |  | 0.77 | 153.5(13.7,293.3) |  | 0.03 |
| O<sub>3</sub> | 2.5(-25.3,30.3) |  | 0.87 | -4.5(-32.8,23.8) |  | 0.75 | 20.5(-39.3,80.3) |  | 0.50 |

## Notes

### Funding Statement

Funded by the Taishan Scholar Program.

